# Machine Learning Approaches for the Prediction Bone Mineral Density by using genomic and phenotypic data of 5,130 older Men

**DOI:** 10.1101/2020.01.20.20018143

**Authors:** Qing Wu, Fatma Nasoz, Jongyun Jung, Bibek Bhattarai, Mira V Han

## Abstract

**Background:** The study aimed to utilize machine learning (ML) approaches and genomic data to develop the prediction model for bone mineral density (BMD), and to identify the best modeling approach for BMD prediction.

**Method:** The genomic and phenotypic data of Osteoporotic Fractures in Men Study (n=5,130), was analyzed. Genetic risk score (GRS) was calculated from 1,103 associated SNPs for each participant after a comprehensive genotype imputation. Data were normalized and divided into a training set (80%) and a validation set (20%) for analysis. Random forest, gradient boosting, neural network, and linear regression were used to develop prediction models for BMD separately. The 10-fold cross-validation was used for hyperparameter optimization. Mean square error and mean absolute error were used to assess model performance. Results: When using GRS and phenotypic covariates as the predictors, the performance of all ML models and linear regression in BMD prediction is similar. However, when replacing GRS with the 1,103 individual SNPs in the model, ML models performed significantly better than linear regression, and the gradient boosting model performed the best. Conclusion: Our study suggested that ML models, especially gradient boosting, can improve BMD prediction in genomic data.

## Introduction

Osteoporosis is a major bone disease characterized by reduced bone mineral density (BMD) and deteriorated bone architecture, leading to increased fracture risk. Osteoporosis and its major complication, osteoporotic fracture, which affects both men and women, cause substantial morbidity and mortality worldwide [1]. Although women have a higher risk of osteoporosis, men suffer much higher morbidity and mortality rates following osteoporotic fractures, especially at an advanced age. With populations aging worldwide, osteoporosis has become a critical public health problem globally. The worldwide fracture incidence in hip alone is projected to increase by 310% in men and 240% in women by 2050, compared to rates in 1990 [2]. The potentially high cumulative rate of fracture, which often results in excess disability and mortality [3], has caused an inevitable increase in the social and economic burden associated with bone health.

BMD has remained the operational definition of osteoporosis since 1994. Osteoporosis is defined by the World Health Organization as a BMD that lies 2.5 or more standard deviations below the average value for young, healthy women. BMD is the single strongest predictor of primary osteoporotic fracture [4]. Each standard deviation decrease in BMD is associated with a 1.5-3.0 fold increase in the risk of fracture, depending on the skeletal region measured, type of fracture, and ethnicity of the study population [5].

BMD is a highly heritable trait. Genetic differences in BMD are well documented [6]. Family and twin studies show BMD variances of 50-85% are attributable to genetic factors [7]. Other studies report BMD heritability estimates of 72-92 [8]. In the past decade, major genome-wide association studies (GWAS) and genome-wide meta-analyses have successfully identified numerous BMD-associated Single Nucleotide Polymorphisms (SNPs) associated with decreased BMD [9]. However, combining these large number of highly significant SNPs, surprisingly, only explained a very small percentage of BMD variance [10]. Such inconsistency may be caused by limitations of the conventional regression approaches employed as these traditional approaches lack the flexibility and adequacy to model complex interactions and regulations.

Linear regression has been widely used as the conventional approach to predicting the BMD outcome [11]. Machine learning (ML) focuses on implementing computer algorithms capable of maximizing predictive accuracy from complex data. ML has a much better capacity to model complex real-world relationships, including variable interactions. Several ML techniques have been applied in clinical research for disease prediction, and ML has shown much higher accuracy for diagnosis than conventional methods [12]. Gradient boosting, random forest, and neural network are widely used ML approach for modeling complex medical data [12]. However, the performance of these ML models for BMD prediction remains unknown, especially with genomic data.

Hence, the aims of the current study are 1) to develop models using ML algorithms to predict BMD from the data with genomic variants, and 2) to compare these models to determine which ML model performs the best for BMD prediction. We hypothesize that when we utilize the ML models to predict BMD, ML models will perform better than linear regression.

## Materials and Methods

### Data Source

The Osteoporotic Fractures in Men Study (MrOS) was used as the data source for this study. MrOS is a federal funded prospective cohort study that was designed to investigate anthropometric, lifestyle, and medical factors associated with bone health in older, community-dwelling men. Details of the MrOS study design, recruitment, and baseline cohort characteristics have been reported [13] elsewhere. With the approval of the institutional review board at the University of Nevada, Las Vegas, and National Institute of Health (NIH), the genotype and phenotype data of MrOS were acquired from dbGaP (Accession: phs000373.v1.p1). MrOS consisted of 5,130 subjects, all of whom had both genotype and phenotype data available for authorized access.

### Study participants

Participants in the MrOS were at least 65 years old, community-dwelling, ambulatory, and had not received bilateral hip replacement [14] at the study entry. At enrollment, participants had to provide self-reported data, understand and sign the written informed consent, complete the self-administered questionnaire, attend a clinic visit, and complete at least the anthropometric, DEXA, and vertebral X-ray procedures. The participants could not have a medical condition that would result in imminent death, which was based on the judgment of the investigators. A total of 5,994 men were enrolled between March 2000 and April 2002, all from six communities in the United States (Birmingham, AL; Minneapolis, MN; Palo Alto, CA; Pittsburgh, PA; Portland, OR; and San Diego, CA.) [15].

### Outcome BMD measurements

Total body, total femur BMD, and lumbar spine (L1 to L4) were measured using a fan-beam dual-energy X-ray absorptiometry (QDR 4500 W, Hologic, Inc., Bedford, MA, USA) at the second visit of MrOS. Participants were scanned for BMD measurements by licensed densitometrists using standardized procedures. All DXA operators were centrally certified based on the evaluation results of scanning and analysis techniques. Cross-calibrations, which were conducted prior to participants’ visits for BMD measurement, found no linear differences across scanners, and the maximum percentage difference between scanners was 1.4% in mean BMD of the total spine [16]. No shifts or drifts in scanner performance was found, based on daily quality control in each clinical center.

### Assessment of covariates

Bone health-related information, including demographics, clinical history, medications, and lifestyle factors, were obtained by self-administered questionnaires. The information collected contained the variables used in this study, including age, race, smoking, and alcohol consumption. Height (cm) was measured using a Harpenden stadiometer, and weight (kg) was measured by a standard balance beam or an electric scale.

Smoking was categorized as “never,” “past,” and “current.” Alcohol intake was quantified in terms of the usual number of drinks per day. Walking speed was determined by timed completion of a 6-meter course, performed at each participant’s typical walking speed. Mobility limitations were quantified by using a participant’s ability to rise from a chair without using his arms, as well as his ability to complete five chair stands. Each participant’s function status was quantified by assessing the difficulty of daily living on a scale of 0-3, with five instrumental activities of daily living, which include walking on level ground, climbing steps, preparing meals, performing housework, and shopping.

### Genotyping Data

Whole blood samples at the baseline were used for DNA extraction. Consent for DNA use was obtained through written permission. Quality-control genotype data files were acquired through dbGaP. Genotype imputation was conducted at the Sanger Imputation Server. The Haplotype Reference Consortium imputation reference panel, and Positional Burrows-Wheeler Transform imputing algorithm, were used to ensure high quality of genotype imputation. Based on the study published by Morris et al. in 2019, a total of 1,103 associated SNPs were extracted for this analysis [9]. All the 1,103 SNPs were successfully imputed in the MrOS data and were included in the analysis. The imputation quality was excellent, with a mean R2 of 0.99. Genotyping for MrOS samples was performed with the Illumina HumanOmni1_Quad_v1-0 H array. A total of 934,940 SNP markers with known chromosome locations, and SNP markers with minor allele frequencies greater than or equal to 0.05, were analyzed.

### Genetic risk score

A genetic risk score (GRS) is a standardized metric that allows the composite assessment of genetic risk in complex traits. The GRS was derived from the number of risk alleles and their effect size for each study subject. We performed a linkage disequilibrium (LD) pruning in advance in order to eliminate possible LD between SNPs; however, none of the SNPs were eligible for removal during the pruning process. The weighted GRS was then calculated with the algorithms described previously [17]. Briefly, for each participant in MrOS, weighted GRS was calculated by summing the number of risk alleles at each locus weighted by regression coefficients related to BMD [9].

### Data processing

Figure 1 shows an overview of our data process flow for this study. After genotype imputation, the phenotype data set (n=5,143) and genotype data set (n=5,130) were merged, and 13 participants were removed from the analysis due to the lack of all phenotype data. We normalized all continuous variables in the data, then randomly divided the dataset into a training set (80%, n=4,104) and a test set (20%, n=1,026). The median imputation [18], the most common imputation method for continuous variables, was used to replace missing values in the data so as to maximize the sample size for analysis.

**Figure 1.**
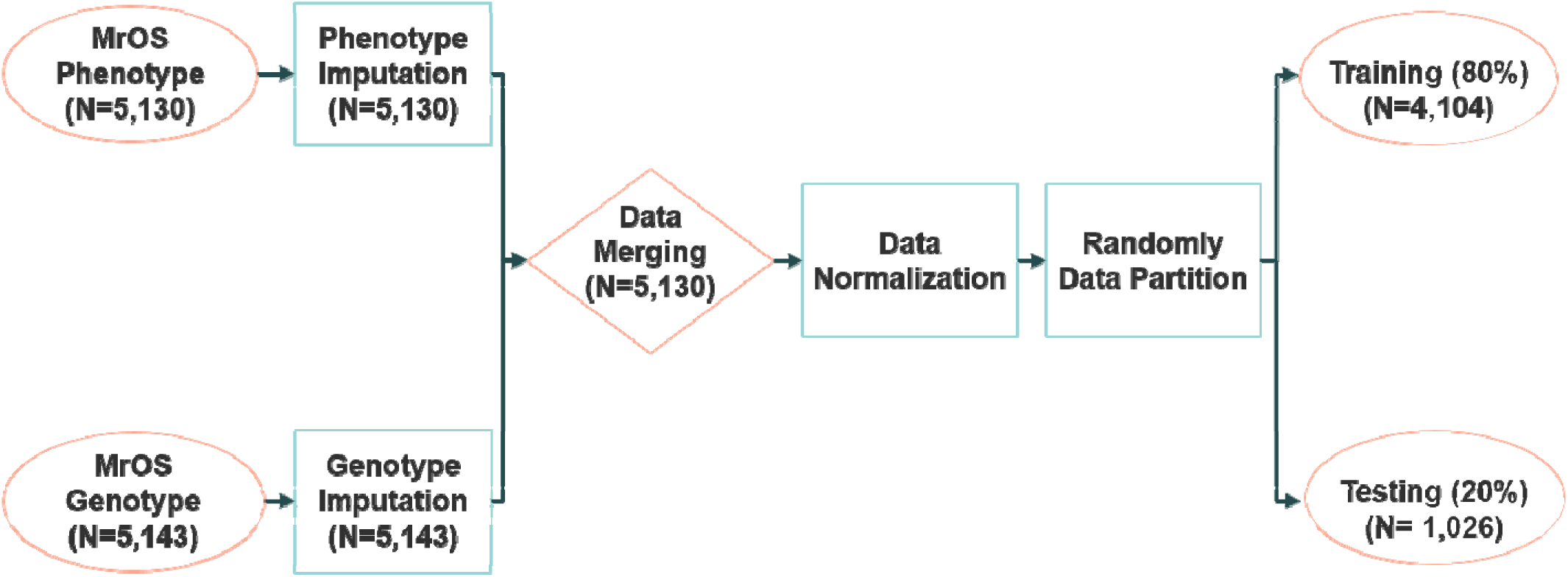
Overview of data process flow.

### Data analysis

The outcomes variables were BMD measured from various skeletal regions, which included femoral neck, total spine, and total hip. The predictors included GRS, age, race, body weight, height, smoking, alcohol consumption, walking speed, impairment of instrumental activities of daily living, mobility limitations. Linear regression, random forest, gradient boosting, and neural network with backpropagation were used to train the model separately. We also conducted analyses that replaced the GRS with the 1,103 individual SNPs in each model. We encoded each risk SNP as three different genotypes (dominant homozygous allele, heterozygotes, homozygous minor allele) with 0, 1, and 2, respectively.

In model training, 10-fold cross-validation was used for hyper-parameter optimization. We divided the training set into 10-folds, and chose one fold as a validation set, with the remaining folds used as the training set. We used the method of Scikit-learn’s randomized search cross-validation [19] to find the best hyperparameters for different algorithms. The training set was used to train and construct the models of linear regression, random forest, gradient boosting, and neural network. A small learning rate and relatively small depth were used for random forest and gradient boosting algorithms in order to reduce the risk of overfitting [20]. With phenotype covariate and GRS as predictors, we used the depth of three for total hip BMD, the depth of two for femur neck BMD, and the depth of one for total spine BMD. With phenotype covariate and 1,103 individual SNPs as predictors, we used the depth of four for total hip BMD, the depth of three at femoral neck BMD, and the depth of one for total spine BMD. In the neural network model, with individual SNPs as predictors, Lasso was used to address the overfitting problem [21].

The testing set (20%) was used to evaluate the prediction performance of the developed model. Metrics for model performance evaluation are mean squared error and mean absolute error [22]. We adopted both metrics:

- Mean Squared Error (MSE)

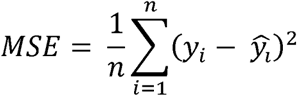
- Mean Absolute Error (MAE)

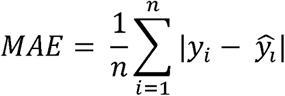

where *n* is the sample size, *y_i_* is the actual value for each observation, and 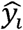 is the estimated value for each observation from the model. We first used MSE as a loss function to develop the model in a training set. We also calculated MSE for each model in the test set and used the MSE for model evaluation. We then reanalyzed the data by replacing MSE with MAE, in which MAE was used as a loss function to develop the model in a training set, and was calculated in the test set for model evaluation. Wilcoxon signed-rank test was employed to examine the difference of MSE or MAE between ML models, as the data distribution assumption for the student t-test was not met. All of the analyses were performed in the Python Software Foundation and Python Language Reference, version 3.7.3, with the package Scikit-learn: Machine Learning in Python [19].

## Results

### Baseline characteristics

Table 1 shows the characteristics of participants within the training (***n*** = **4,104**) and the test (***n*** = **1,026**) datasets. Demographic and clinical variables were not significantly different in training and test datasets. All BMD measurements from the femur neck, total spine, and total hip were normally distributed, with means 0.78, 0.96, 1.07, and standard deviations 0.13, 0.14, 0.19, respectively.

**Table 1.** Baseline characteristics of training and testing dataset.

| Variable* | Training<br>Dataset<br>(*n* = 4,104) | Test<br>Dataset<br>(*n* = 1,026) | *p* – value** |
| --- | --- | --- | --- |
| Femoral Neck BMD<br>(g/cm<sup>2</sup>) | 0.78 ± 0.13 | 0.79 ± 0.13 | 0.61 |
| Total Hip BMD<br>(g/cm<sup>2</sup>) | 0.96 ± 0.14 | 0.96 ± 0.14 | 0.55 |
| Total Spine BMD<br>(g/cm<sup>2</sup>) | 1.07 ± 0.19 | 1.07 ± 0.19 | 0.36 |
| Age (year) | 73.77 ± 5.93 | 73.95 ± 5.83 | 0.78 |
| Height (cm) | 174.15 ± 6.81 | 174.16 ± 6.61 | 0.69 |
| Weight (kg) | 83.28 ± 13.46 | 82.73 ± 12.73 | 0.37 |
| Alcohol use | 4.25 ± 6.87 | 3.98 ± 6.06 | 0.57 |
| GRS*** | 31.6 ± 0.44 | 31.59 ± 0.45 | 0.81 |
| Impairment of<br>Instrumental<br>Activities of Daily Living | 0.37 ± 0.88 | 0.36 ± 0.83 | 0.79 |
| Walking Speed | 1.07 ± 0.27 | 1.08 ± 0.29 | 0.53 |
| Smoking, No. (%) |  |  |  |
| No | 1,535 (37.4%) | 406 (39.6%) | 0.41 |
| Past | 2,430 (59.2%) | 587 (57.3%) |  |
| Current | 139 (3.4%) | 32 (3.1%) |  |
| Race, No. (%) |  |  |  |
| White | 3,707 (90.3%) | 909 (88.6%) | 0.11 |
| African American | 141 (3.4%) | 40 (3.9%) |  |
| Asian | 120 (2.9%) | 45 (4.4%) |  |
| Hispanic | 87 (2.1%) | 24 (2.3%) |  |
| Other | 49 (1.2%) | 8 (0.8%) |  |

\* Continuous variables were expressed as mean ± SD, and categorical variables were expressed as number (%).
\*\* *p*- values were obtained by *t* – test for continuous variables and chi-square tests for the categorical variable.
\*\*\* GRS: genetic risk score, which was calculated based on 1,103 BMD-related SNPs.

### Model Performance

Figure 2 shows the performance of each model in the test dataset (***n*** = **1,026**). The upper panel in Figure 2 compared model performance when using phenotype covariates and GRS as predictors. MSE in each model became similar in the test data with increased training iterations. Although the linear regression model had a relatively higher MSE in the first few iterations, the performance of linear regression and ML models became nearly identical after 100 iterations of training for BMD in each skeletal region. All models had the best performance in femur neck BMD (Figure 2A) with the lowest MSE during training iteration, followed by total hip BMD (Figure 2B), and then total spine BMD (Figure 2C). The lower panel in Figure 2 compared model performance when using phenotype covariates and 1,103 individual SNPs as predictors. MSE in linear regression was much larger than that in other ML models in the test data, even after 100 iterations of training; the results were consistent with BMD measured at the three different skeletal regions.

**Figure 2.**
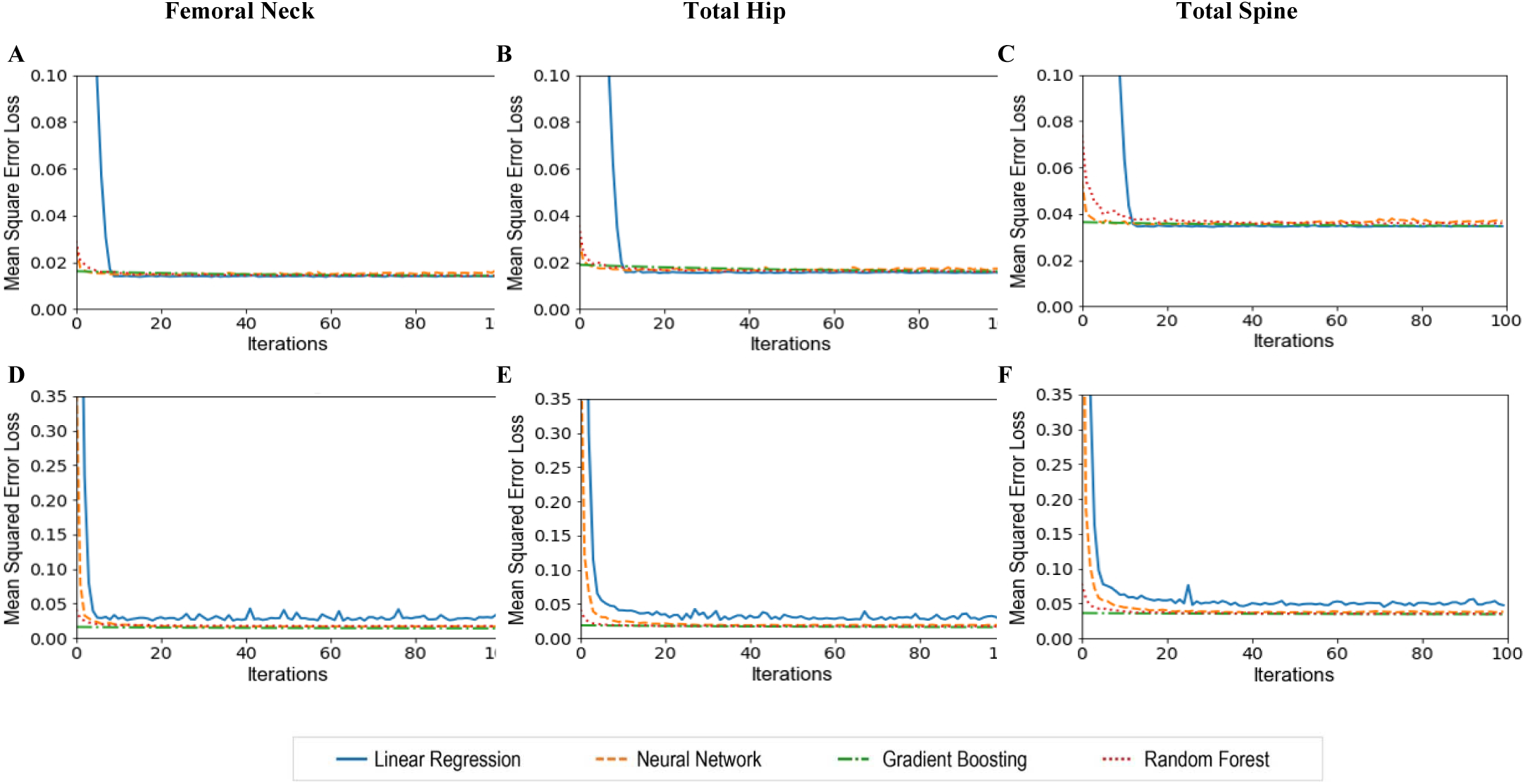
Mean squared error loss of various models with the number of training iterations for BMD prediction in the test dataset (**n = 1,026**). The upper panel shows the performance of each model with phenotype covariates and GRS as predictors in predicting BMD at the femoral neck (A), total hip (B), and total spine (C) in the testing dataset at different BMD sites. The lower panel shows the performance of each model with phenotype covariates and 1,103 individual SNPs in predicting BMD at the femoral neck (D), total hip (D), and total spine (F).

Tables 2 show the results of MSE and MAE for each model with phenotype covariate and GRS as predictors. In the test dataset (***n*** = **1,026**), MSEs were similar between all models in each BMD, and the same results were observed for MAE. When we replaced GRS with 1,103 SNPs in the predictors (Table 3), in the testing dataset, ML models had smaller MSE than that in the linear model, and similar results were observed with MAE. Overall the gradient boosting model had the lowest MSE and MAE in the testing dataset.

**Table 2.** Mean Square Error (MSE) and Mean Absolute Error (MAE) of different models in predicting various BMD in the test dataset (*n* = 1,026); GRS and phenotypic covariates were used as predictors.

|  | MSE |  | MAE |  |
| --- | --- | --- | --- | --- |
|  | Testing | Training | Testing | Training |
| **Femoral Neck BMD** |  |  |  |  |
| **Linear Regression** | 0.0146 | 0.0126 | 0.0941 | 0.0881 |
| **Random Forest** | 0.0152 | 0.0151 | 0.0945 | 0.0960 |
| **Gradient Boosting** | 0.0142 | 0.0141 | 0.0946 | 0.0360 |
| **Neural Network** | 0.0150 | 0.0122 | 0.0958 | 0.0868 |
| **Total Hip BMD** |  |  |  |  |
| **Linear Regression** | 0.0164 | 0.0151 | 0.1008 | 0.0967 |
| **Random Forest** | 0.0163 | 0.0163 | 0.1003 | 0.1005 |
| **Gradient Boosting** | 0.0162 | 0.0157 | 0.1026 | 0.0394 |
| **Neural Network** | 0.0162 | 0.0153 | 0.1005 | 0.0974 |
| **Total Spine BMD** |  |  |  |  |
| **Linear Regression** | 0.0365 | 0.0316 | 0.1489 | 0.1363 |
| **Random Forest** | 0.0353 | 0.0353 | 0.1469 | 0.1439 |
| **Gradient Boosting** | 0.0349 | 0.0347 | 0.1490 | 0.0562 |
| **Neural Network** | 0.0352 | 0.0324 | 0.1460 | 0.1378 |

**Table 3.**
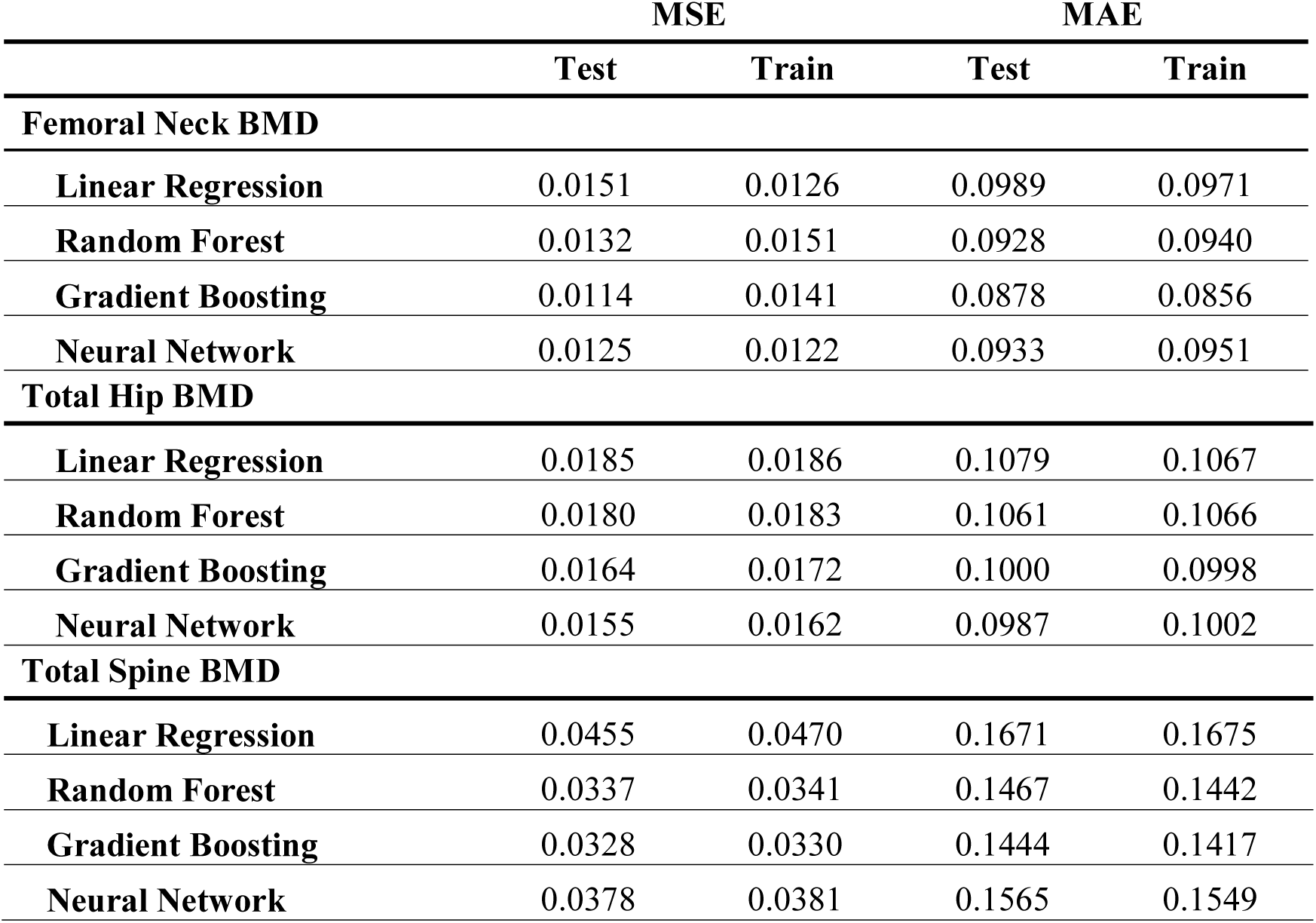
Mean Square Error (MSE) and Mean Absolute Error (MAE) of different models in predicting various BMD in the test dataset (*n* = 1,026); 1,103 SNPs and phenotypic covariates were used as predictors.

|  | MSE |  | MAE |  |
| --- | --- | --- | --- | --- |
|  | Test | Train | Test | Train |
| **Femoral Neck BMD** |  |  |  |  |
| **Linear Regression** | 0.0151 | 0.0126 | 0.0989 | 0.0971 |
| **Random Forest** | 0.0132 | 0.0151 | 0.0928 | 0.0940 |
| **Gradient Boosting** | 0.0114 | 0.0141 | 0.0878 | 0.0856 |
| **Neural Network** | 0.0125 | 0.0122 | 0.0933 | 0.0951 |
| **Total Hip BMD** |  |  |  |  |
| **Linear Regression** | 0.0185 | 0.0186 | 0.1079 | 0.1067 |
| **Random Forest** | 0.0180 | 0.0183 | 0.1061 | 0.1066 |
| **Gradient Boosting** | 0.0164 | 0.0172 | 0.1000 | 0.0998 |
| **Neural Network** | 0.0155 | 0.0162 | 0.0987 | 0.1002 |
| **Total Spine BMD** |  |  |  |  |
| **Linear Regression** | 0.0455 | 0.0470 | 0.1671 | 0.1675 |
| **Random Forest** | 0.0337 | 0.0341 | 0.1467 | 0.1442 |
| **Gradient Boosting** | 0.0328 | 0.0330 | 0.1444 | 0.1417 |
| **Neural Network** | 0.0378 | 0.0381 | 0.1565 | 0.1549 |

The nonparametric Wilcoxon signed-rank test results for multiple comparisons of MSE between models are shown in Table 4; when using phenotype covariates and GRS as the predictors. With Bonferroni corrections for multiple comparisons (α = 0.05/6=0.0083), none of the comparisons were statistically significant except the comparison between gradient boosting and random forest at femur neck BMD and total spine BMD. However, as shown in Table 5, when using phenotype covariates and 1,103 SNPs as the predictors, the difference of MSE in most pairwise comparisons was statistically significant with p< .0001. The only exceptions are the comparison between neural network and random forest at femur neck BMD and the comparison between gradient boosting and random forest at total hip BMD with p> .05.

**Table 4.** Statistical comparisons of mean square errors in the testing dataset (*n* = 1,026) between various models when phenotype covariates and GRS were used as the predictors. The Wilcoxon Signed-Rank Test was used to determine all p-values.

|  | Linear Regression | Random Forest | Gradient Boosting |
| --- | --- | --- | --- |
| **Femoral Neck BMD** |  |  |  |
| Neural Network | > .05 | > .05 | > .05 |
| Gradient Boosting | > .05 | < .0001 | -- |
| Random Forest | > .05 | -- | -- |
| **Total Hip BMD** |  |  |  |
| Neural Network | > .05 | > .05 | > .05 |
| Gradient Boosting | > .05 | > .05 | -- |
| Random Forest | > .05 | -- | -- |
| **Total Spine BMD** |  |  |  |
| Neural Network | > .05 | > .05 | > .05 |
| Gradient Boosting | > .05 | < .01 | -- |
| Random Forest | > .05 | -- | -- |

**Table 5.** Statistical comparisons of mean square errors in the testing dataset (*n* = 1,026) between various models when phenotype covariates and 1,103 SNPs were used as the predictors. The Wilcoxon Signed-Rank Test was used to determine all p-values.

|  | Linear Regression | Random Forest | Gradient Boosting |
| --- | --- | --- | --- |
| **Femoral Neck BMD** |  |  |  |
| Neural Network | < .0001 | > .05 | < .0001 |
| Gradient Boosting | < .0001 | < .0001 | -- |
| Random Forest | < .0001 | -- | -- |
| **Total Hip BMD** |  |  |  |
| Neural Network | < .0001 | < .0001 | < .0001 |
| Gradient Boosting | < .0001 | > .05 | -- |
| Random Forest | < .0001 | -- | -- |
| **Total Spine BMD** |  |  |  |
| Neural Network | < .0001 | < .0001 | < .0001 |
| Gradient Boosting | < .0001 | < .001 | -- |
| Random Forest | < .0001 | -- | -- |

## Discussion

This study presented findings that employing various ML models and linear regression in BMD prediction in older men, by using both genotype and phenotype data. Interestingly, we found that if we use GRS, the summarized genetic risk from associated SNPs, as the genetic predictor in the model, all ML approaches did not perform better than linear regression in predicting BMD. In contrast, if we replace GRS with the 1,103 individual risk SNPs as predictors in the model, ML models all have significantly better performance than linear regression for BMD prediction. With the increasing availability of genomic and health big data, ML technologies, which employ a wide-ranging class of algorithms, have increasingly been utilized in medical research, especially in disease prediction using genomic and health big data. However, our study findings suggested that the conventional approach may be sufficient if we use GRS, a summary metric for genetic risk, as the genetic predictor in the prediction model. ML approaches are recommended if a large number of individual genetic variants are included as predictors.

ML models have been used widely for prediction in classification problems, especially for disease prediction. However, studies that utilized ML technologies to predict the quantitative trait are still few. Reportedly, artificial neural networks were utilized for BMD prediction in a small sample of Japan postmenopausal women by using common risk factors and a BMD previously measured [11]. However, to the best of our knowledge, the present study is the first attempt to predict BMD using both advanced ML approaches and genomic information, as well as the first to identify the best ML model for BMD prediction. Our study demonstrated ML technologies perform better than conventional methods for the prediction of quantitative traits in complex data that include a large number of genomic variants as predictors.

Risk SNPs, identified in GWAS and Genome-Wide meta-analyses, have posed a challenge in conventional statistical analysis because their effect size is so small. Each associated SNP contributes minimally to the variance of BMD. Thus, GRS is widely used to integrate the effects of individual associated SNPs into a single genetic summary variable for the prediction research in many studies. Although such an approach improves the prediction ability, many uncertainties remain. For example, this approach does not account for gene interaction and regulation. To address these limitations, we utilized ML approaches in the current study so that individual SNPs can be included to replace GRS in the modeling process. ML approaches are able to incorporate the various nonlinear interactions between genetic variants/predictors, which cannot be addressed by conventional modeling methods. Thus, ML approaches provide great potential for improving BMD prediction. In the present study, we employed random forest, gradient boosting, and neural networks, as well as 1,103 individual related SNPs, to find a more accurate BMD prediction model. We found that the gradient boosting model performs best in predicting BMD as it has the lowest MSE and MAE in the validation for all three BMD outcomes. The highest predictive performance of the gradient boosting model has been used widely in predicting various diseases and outcomes, including hip fractures [23]], sepsis [24], urinary tract infections [25], hepatocellular carcinoma [26], and bioactive molecules [27]. The present study suggested that the gradient boosting approach, combined with individual SNPs as the predictors, can provide a more accurate prediction for BMD.

Our study has several strengths. To ensure our study results are robust, first, we have used two metrics, MSE and MAE, to examine and compare the prediction accuracy of the four models we developed. The study findings are consistent between analyses using the two metrics; second, we performed data analysis separately for the outcome variable BMD measured in three different skeletal regions, and the study results were consistent. Finally, we employed the nonparametric Wilcoxon signed-rank test to examine the significance of the difference MSE (and MAE) between any two models in order to ensure the data distribution did not bias the results of statistical tests. We also used Bonferroni corrections for the multiple comparisons to ensure our conclusions were robust.

However, the study has some limitations as well. First, the study sample size (***n*** = **5,130**) is relatively small for ML methods. ML methods often require a much larger sample size for training. To address this limitation, we used 10-fold cross-validation for tuning of the hyper-parameters within the training dataset, so we do not need to allocate part of the study sample for model validation, thus maximizing sample size for the training model. Second, some covariates were not available in the MrOS through dbGaP, including related medications, comorbidities, and physical activities. Lacking these phenotypic variables can impact the performance of all prediction models. Third, the MrOS data only included men ≥ **65** years old and who were mostly Caucasian (90%), so findings from the present study may not apply to women, younger individuals, or other ethnicities. Finally, rare risk SNPs were less likely to be included for modeling in this study, because risk SNPs used in this study were identified from a GWAS study, which likely discovered common variants, not rare variants [28]. Nevertheless, these limitations are unlikely to have altered our findings in the current study because this is a self-control study, with all models developed and validated by the same datasets.

In summary, there was not a significant difference in predicting BMD between various ML models and linear regression if GRS, a metric used to summarize genetic variants, was used for model development. However, when using a large number of individual SNPs as predictors to replace GRS, ML models performed significantly better than linear regression in BMD prediction. Among these ML models, the gradient boost model performed best for BMD prediction. Our study suggests that ML models, especially gradient boosting, can be used to identify patients with low BMD if their genetic information is available. Our study also suggested when researchers used a large number of genetic variants or other predictors, ML approaches, especially gradient boosting, should be considered. Additional, more comprehensive studies, especially those including women, young participants, rare genetic variants, and additional risk factors, are warranted to further examine the research findings from the present study.

## Data Availability

The Osteoporotic Fractures in Men Study (MrOS) was used as the data source for this study. MrOS is a federal funded prospective, cohort study which was designed to investigate anthropometric, lifestyle, and medical factors associated with bone health in older, community‐dwelling men. Details of the MrOS study design, recruitment, and baseline cohort characteristics have been reported12 elsewhere. With the approval of the institutional review board at the University of Nevada, Las Vegas and National Institute of Health (NIH), the genotype and phenotype data of MrOS was acquired from dbGaP (Accession: phs000373.v1.p1). The data/analyses presented in the current publication are based on the use of study data downloaded from the dbGaP web site, under phs000373.v1.p1 (https://www.ncbi.nlm.nih.gov/projects/gap/cgi-bin/study.cgi?study_id=phs000373.v1.p1).

## Author Contributions

Conceptualization: QW; methodology: QW, FN, JJ, and BB; software: JJ and BB; validation: QW, FN, JJ, BB, and MH; formal analysis: JJ and BB; investigation: QW, FN, JJ, BB, and MH; resources: QW; data curation: JJ and BB; writing-original draft preparation: QW, and JJ; writing-review and editing: QW, FN, JJ, and MH; visualization: JJ and BB; supervision, QW; project administration: QW; funding acquisition: QW. All authors have read and agreed to the published version of the manuscript.

## Acknowledgments

The data/analyses presented in the current publication are based on the use of study data downloaded from the dbGaP web site, under phs000373.v1.p1 (https://www.ncbi.nlm.nih.gov/projects/gap/cgi-bin/study.cgi?study_id=phs000373.v1.p1). The research and analysis described in the current publication were supported by the Genome Acquisition and Analytics (GAA) Research Core of the Personalized Medicine Center of Biomedical Research Excellence in the Nevada Institute of Personalized Medicine. The National Supercomputing Institute at the University of Nevada Las Vegas provided facilities for bioinformatical analysis in this study.

## Abbreviations

MrOS: :Osteoporotic Fractures in Men Study
BMD: :Bone Mineral Density
ML: :Machine Learning
GRS: :Generic Risk Score
LR: :Linear Regression
RF: :Random Forest
GB: :Gradient Boosting
NN: :Neural Network
SNPs: :Single Nucleotide Polymorphisms
GWAS: :Genome-Wide Association Study

## References

1. Cummings, S.R.; Melton, L.J. Epidemiology and outcomes of osteoporotic fractures. Lancet 2002, 359, 1761–1767, doi:10.1016/S0140-6736(02)08657-9.

2. Gullberg, B.; Johnell, O.; Kanis, J.A. World-wide projections for hip fracture. Osteoporos Int 1997, 7, 407–413, doi:10.1007/PL00004148.

3. Melton, L.J.; Cooper, C. Chapter 21 - Magnitude and Impact of Osteoporosis and Fractures. In Osteoporosis; Academic Press Inc., 2007; pp. 557–567.

4. Kanis, J.A.; Borgstrom, F.; De Laet, C.; Johansson, H.; Johnell, O.; Jonsson, B.; Oden, A.; Zethraeus, N.; Pfleger, B.; Khaltaev, N. Assessment of fracture risk. Osteoporos Int 2005, 16, 581–589, doi:10.1007/s00198-004-1780-5.

5. Marshall, D.; Wedel, H. Meta-analysis of how well measures of bone mineral density predict occurrence of osteoporotic fractures. theBMJ 1996, 312, 1254–1259, doi:10.1136/bmj.312.7041.1254.

6. Warrington, N.M.; Kemp, J.P.; Tilling, K.; Tobias, J.H.; Evans, D.M. Genetic variants in adult bone mineral density and fracture risk genes are associated with the rate of bone mineral density acquisition in adolescence. Hum Mol Genet 2015, 24, 4158–4166, doi:10.1093/hmg/ddv143.

7. Eisman, J.A. Genetics of Osteoporosis. Endocr Rev 1999, 20, 788–804, doi:10.1002/9780470623992.ch42.

8. Nicholas A. Pocock, John A. Eisman, John L. Hopper, Michael G. Yeates, Philip N. Sambrook, S.E. Genetic determinants of bone mass in adults. A twin study. Jounral Clin Investig 1987, 80, 706–710.

9. Morris, J.A.; Kemp, J.P.; Youlten, S.E.; Laurent, L.; Logan, J.G.; Chai, R.C.; Vulpescu, N.A.; Forgetta, V.; Kleinman, A.; Mohanty, S.T.; et al. An atlas of genetic influences on osteoporosis in humans and mice. Nat Genet 2019, 51, 258–266, doi: 10.1038/s41588-018-0302-x.

10. Xiao, X.; Roohani, D.; Wu, Q. Genetic profiling of decreased bone mineral density in an independent sample of Caucasian women. Osteoporos Int 2018, 29, 1807–1814, doi: 10.1007/s00198-018-4546-1.

11. Shioji, M.; Yamamoto, T.; Ibata, T.; Tsuda, T.; Adachi, K.; Yoshimura, N. Artificial neural networks to predict future bone mineral density and bone loss rate in Japanese postmenopausal women. BMC Res Notes 2017, 10, 1–5, doi: 10.1186/s13104-017-2910-4.

12. Hsieh, C.H.; Lu, R.H.; Lee, N.H.; Chiu, W.T.; Hsu, M.H.; Li, Y.C. Novel solutions for an old disease: Diagnosis of acute appendicitis with random forest, support vector machines, and artificial neural networks. Surgery2011, 149, 87–93, doi:10.1016/j.surg.2010.03.023.

13. Orwoll, E.; Blank, J.B.; Barrett-Connor, E.; Cauley, J.; Cummings, S.; Ensrud, K.; Lewis, C.; Cawthon, P.M.; Marcus, R.; Marshall, L.M.; et al. Design and baseline characteristics of the osteoporotic fractures in men (MrOS) study - A large observational study of the determinants of fracture in older men. Contemp Clin Trials 2005, 26, 569–585, doi:10.1016/j.cct.2005.05.006.

14. Riggs, L.; Melton, L. The worldwide problem of osteoporosis: lessons from epidemiology. Bone 1995, 17, 2–3.

15. Blank, J.B.; Cawthon, P.M.; Carrion-Petersen, M. Lou; Harper, L.; Johnson, J.P.; Mitson, E.; Delay, R.R. Overview of recruitment for the osteoporotic fractures in men study (MrOS). Contemp Clin Trials 2005, 26, 557–568, doi:10.1016/j.cct.2005.05.005.

16. Cauley, J.A.; Fullman, R.L.; Stone, K.L.; Zmuda, J.M.; Bauer, D.C.; Barrett-Connor, E.; Ensrud, K.; Lau, E.M.C.; Orwoll, E.S. Factors associated with the lumbar spine and proximal femur bone mineral density in older men. Osteoporos Int 2005, 16, 1525–1537, doi:10.1007/s00198-005-1866-8.

17. Andrews, N.A. Genome-wide association studies in the osteoporosis field: Impressive technological achievements, but an uncertain future in the clinical setting. IBMS Bonekey 2010, 7, 382–387, doi:10.1138/20100472.

18. Gao, B. Advances in Intelligent Systems and Computing 2019; Vol. 997; ISBN 978-3-030-01173-4.

19. Fabian Pedregosa, Gael Varoquaux, Alexandre Gramfort, Vincent Michel, Bertran Thirion, Olivier Grisel, Mathieu Blondel, Peter Prettenhofer, Ron Weiss, Vincent Dubourg, Jake Vanderplas, Alexandre Passos, D.C. Scikit-learn: machine learning in Python. J Mach Learn Res 2011, 12, 2825–2830.

20. Friedman, J.; Hastie, T.; Tibshirani, R. Additive logistic regression: a statistical view of boosting (With discussion and a rejoinder by the authors). Ann Stat 2000, 28, 337–407, doi:10.1214/aos/1016218223.

21. Tibshirani, R. Regression Shrinkage and Selection Via the Lasso. J R Stat Soc Ser B 1996, 58, 267–288, doi:10.1111/j.2517-6161.1996.tb02080.x.

22. Amoroso, N.; Rocca, M. La; Bellantuono, L.; Diacono, D.; Fanizzi, A.; Lella, E.; Lombardi, A.; Maggipinto, T.; Monaco, A.; Tangaro, S.; et al. Deep learning and multiplex networks for accurate modeling of brain age. Front Aging Neurosci 2019, 11, 1–12, doi:10.3389/fnagi.2019.00115.

23. Kruse, C.; Eiken, P.; Vestergaard, P. Machine Learning Principles Can Improve Hip Fracture Prediction. Calcif Tissue Int 2017, 100, 348–360, doi: 10.1007/s00223-017-0238-7.

24. Chiew, C.J.; Liu, N.; Tagami, T.; Wong, T.H.; Koh, Z.X.; Ong, M.E.H. Heart rate variability based machine learning models for risk prediction of suspected sepsis patients in the emergency department. Medicine (Baltimore) 2019, 98, e14197, doi:10.1097/MD.0000000000014197.

25. Taylor, R.A.; Moore, C.L.; Cheung, K.H.; Brandt, C. Predicting urinary tract infections in the emergency department with machine learning. PLoS One 2018, 13, 1–15, doi:10.1371/journal.pone.0194085.

26. Sato, M.; Morimoto, K.; Kajihara, S.; Tateishi, R.; Shiina, S.; Koike, K.; Yatomi, Y. Machine-learning Approach for the Development of a Novel Predictive Model for the Diagnosis of Hepatocellular Carcinoma. Sci Rep 2019, 9, 1–7, doi: 10.1038/s41598-019-44022-8.

27. Babajide Mustapha, I.; Saeed, F. Bioactive Molecule Prediction Using Extreme Gradient Boosting. Molecules 2016, 21, 1–11, doi:10.3390/molecules21080983.

28. Nguyen, T. V.; Eisman, J.A. Genetic profiling and individualized assessment of fracture risk. Nat Rev Endocrinol 2013, 9, 153–161, doi:10.1038/nrendo.2013.3.

